# The impact of SARS-CoV-2 vaccination on Alpha & Delta variant transmission

**DOI:** 10.1101/2021.09.28.21264260

**Authors:** David W Eyre, Donald Taylor, Mark Purver, David Chapman, Tom Fowler, Koen B Pouwels, A Sarah Walker, Tim EA Peto

## Abstract

**Background:** Pre-Delta, vaccination reduced SARS-CoV-2 transmission from individuals infected despite vaccination, potentially via reducing viral loads. While vaccination still lowers the risk of infection, similar viral loads in vaccinated and unvaccinated individuals infected with Delta question how much vaccination prevents transmission.

**Methods:** We performed a retrospective observational cohort study of adult contacts of SARS-CoV-2-infected adult index cases using English contact testing data. We used multivariable Poisson regression to investigate associations between transmission and index case and contact vaccination, and how these vary with Alpha and Delta variants (classified using S-gene detection/calendar trends) and time since second vaccination.

**Results:** 54,667/146,243(37.4%) PCR-tested contacts of 108,498 index cases were PCR-positive. Two doses of BNT162b2 or ChAdOx1 vaccines in Alpha index cases were independently associated with reduced PCR-positivity in contacts (aRR, adjusted rate ratio vs. unvaccinated=0.32[95%CI 0.21-0.48] and 0.48[0.30-0.78] respectively). The Delta variant attenuated vaccine-associated reductions in transmission: two BNT162b2 doses reduced Delta transmission (aRR=0.50[0.39-0.65]), more than ChAdOx1 (aRR=0.76[0.70-0.82]). Variation in Ct values (indicative of viral load) explained 7-23% of vaccine-associated transmission reductions. Transmission reductions declined over time post-second vaccination, for Delta reaching similar levels to unvaccinated individuals by 12 weeks for ChAdOx1 and attenuating substantially for BNT162b2. Protection in contacts also declined in the 3 months post-second vaccination.

**Conclusions:** Vaccination reduces transmission of Delta, but by less than the Alpha variant. The impact of vaccination decreased over time. Factors other than PCR Ct values at diagnosis are important in understanding vaccine-associated transmission reductions. Booster vaccinations may help control transmission together with preventing infections.

## Introduction

SARS-CoV-2 vaccines have been shown in randomised controlled trials^1–3^ and real-world population studies^4,5^ to prevent infection and adverse outcomes from several SARS-CoV-2 variants including Alpha (B.1.1.7) and Delta (B.1.617.2).^6–8^

Vaccination also potentially prevents onward transmission by at least two mechanisms: reducing symptomatic and asymptomatic infections and therefore the number of infectious individuals, and secondly via reduced onward spread from those infected despite vaccination. Household studies show vaccination reduces onward transmission of the Alpha variant from those infected despite vaccination.^9–12^ One hypothesised mechanism is lower viral loads observed in post-vaccination Alpha infections^7,13^ vs. unvaccinated individuals, as viral load is associated with the likelihood of infection in contacts.^14,15^

However, Delta viral loads are similar in vaccinated and unvaccinated infected individuals,^8,16^although the duration of viral shedding may be reduced.^17,18^ This questions whether vaccination controls Delta spread as effectively as Alpha, and whether, with increased transmissibility,^19^ this explains the rapid global dissemination of Delta despite rising vaccination coverage.

We use national contact testing data from England to investigate the impact of vaccination on onward transmission of SARS-CoV-2, and how this varies with Alpha and Delta variants and time since second vaccination.

## Methods

### Setting and variants

We performed a retrospective observational cohort study of adult contacts (≥18y) of symptomatic and asymptomatic SARS-CoV-2-infected adult index cases. Data were obtained from the English national contact tracing and testing service, NHS Test and Trace. Contacts (living in the same household or in contact face-to-face, within <1m for ≥1 minute or <2m for ≥15 minutes) were eligible for inclusion if they accessed PCR testing 1-10 days after the index case’s PCR test (typically following symptoms, but also after positive asymptomatic antigen screening); 1-10 days chosen to enrich for contacts where the index case was the most likely source for any infection^15^ (tested in a sensitivity analysis, see Supplement). Only index cases with PCR tests performed by three national “lighthouse” laboratories (Milton Keynes, Alderley Park, Glasgow) were included, using the same standardised workflow and PCR assay (ThermoFisher TaqPath; S-gene, N-gene and ORF1ab targets). Contacts could be tested by any community/hospital laboratory reporting results to NHS Test and Trace. Vaccination status in cases and contacts was obtained from the National Immunisation Management Service (see Supplement).

Contacts of index cases tested between 01-January-2021 and 31-July-2021 were included as follows. Index cases were classified as Alpha (B.1.1.7) variant based on S-gene target failure (SGTF), while this was considered a reliable proxy for Alpha, namely to 06-June-2021 (after which <6% cases had SGTF). From 10-May-2021 national spread of Delta meant >98% sequenced cases were due to Alpha or Delta variants,^19^ such that we used S-gene detection from 10-May-2021 as a proxy for Delta (see Supplement).

We restricted our analysis to contacts undergoing testing, excluding untested contacts, to control as much as possible for biases related to health-seeking behaviour (including differences before/after vaccination), access to testing, and case ascertainment.^20^

### Statistical analysis

We used multivariable Poisson regression to investigate how onward transmission, i.e., SARS-CoV-2 PCR-positive tests in contacts, varied with index case vaccination status: unvaccinated, partially vaccinated (first vaccine date to 13 days after second vaccine), or fully vaccinated (≥14 days after second vaccine), further considering whether vaccination was AstraZeneca ChAdOx1 or Pfizer-BioNTech BNT162b2. We investigated how onward transmission varied with Alpha vs. Delta index cases and whether any effects varied by vaccine via pre-specified interaction terms. We additionally included model terms for time since second BNT162b2 or ChAdOx1 vaccine.

We adjusted for the following covariates: contact event type; index case factors (age, sex, and symptom status); contact factors (age, sex, vaccination status and time since vaccination, as above); local deprivation, local weekly SARS-CoV-2 incidence from national testing data, and calendar time (reflecting temporal changes in behaviour/social distancing, the likelihood of acquisition from a third party, population-wide vaccine uptake, and the percentage of unvaccinated people previously infected)(Table S1). We accounted for non-linearity, interactions and multiple testing (see Supplement).

We refitted models including index case Ct values to investigate the relationship between Ct values (indicative of viral load^21^) and transmission and performed a mediation analysis to investigate whether the effect of index case vaccination status was explained by Ct values at diagnosis (see Supplement).

### Ethics

The study was performed as public health surveillance and NHS Test and Trace program quality assurance, under Section 251 of the NHS Act 2006 with approvals from Public Health England (PHE), the Department of Health and Social Care and NHS Test and Trace. PHE’s Research Ethics and Governance Group (PHE’s Research Ethics Committee) reviewed the study protocol and confirmed compliance with all regulatory requirements. As no regulatory or ethical issues were identified, it was agreed that full ethical review was not needed, and the protocol was approved.

## Results

661,315 adult contacts of 374,115 adult index cases were recorded; 173,460(26.2%) contacts underwent PCR testing between 02-January-2021 and 02-August-2021. 27,217(15.7%) contacts were excluded with incomplete data (see Supplement). Of the remaining 146,243 contacts (108,498 index cases), 54,667(37.4%) tested PCR-positive. The median(IQR)[range] index case and contact ages were 34(24-49)[18-102] and 43(29-54)[18-107] years respectively. 55,354(51%) index cases and 83,206(57%) contacts were female (Table S1-S2 for details by case/contact vaccine status). Contact events were predominantly within households (97,204;66%), but also in household visitors (16,505;11%), at event/activities (16,114;11%) and work/education (16,420;11%).

### Index case vaccination and onward transmission

35,459/76,401(46%) contacts of unvaccinated index cases tested PCR-positive, as did 3,878/11,236(35%) and 7,947/31,039(26%) contacts of partially ChAdOx1 and BNT162b2 vaccinated cases, and 6,067/21,421(28%) and 1,316/6,146(21%) contacts of fully ChAdOx1 and BNT162b2 vaccinated cases. For index cases vaccinated twice with ChAdOx1 or BNT162b2, the median(IQR) days from second vaccine to an Alpha variant PCR-positive test was 27(18.5-43) and 42(26-63), respectively, and 51(35-70) and 90(69-110) for Delta. Dosing intervals for fully-vaccinated index cases were >6 weeks for 14,811/15,083(98%) receiving ChAdOx1 and 3,759/4,233(89%) for BNT162b2.

In a multivariable model (Tables 1, S4, Figures S2-S4), BNT162b2 vaccination in Alpha variant index cases was independently associated with reduced PCR-positivity in contacts; two doses (adjusted rate ratio at 14 days post-second vaccine vs. unvaccinated, aRR=0.32[95%CI 0.21-0.48]) reduced onward transmission more than one (aRR=0.88[0.85-0.91]). Similarly, for ChAdOx1, two doses reduced transmission (aRR=0.48[0.30-0.78]) more than one (aRR=0.90[0.86-0.94]). There was no evidence of a difference in transmission reductions after two doses between vaccines for Alpha (heterogeneity RR, hRR=1.51[0.81-2.85]).

**Table 1.** Relationship between PCR-positive results in contacts, and index case and contact vaccination status according to Alpha/Delta variant in the index case. Results for those with two vaccine doses are estimated at day 14 post second vaccine, see Figure 1 for trends with time post-second vaccine. aRR, adjusted rate ratio, CI confidence interval. Adjustment made for contact event type; index case factors - age, sex, and symptom status; contact factors - age, sex; local deprivation, local SARS-CoV-2 incidence and calendar time (see Table S4 and Figures S2-S4 for details). There was no evidence that adding an interaction between case and contact vaccination status improved model fit.

|  | Alpha |  | Delta |  | Delta vs. Alpha |  |
| --- | --- | --- | --- | --- | --- | --- |
| Characteristic | aRR | 95% CI | aRR | 95% CI | Interaction RR | 95% CI |
| ***Impact on onward transmission: Case vaccination status*** |  |  |  |  |  |  |
| Unvaccinated | — | — | — | — | — | — |
| Partial ChAdOx1 | 0.90 | 0.86, 0.94 | 0.95 | 0.91, 0.99 | 1.06 | 1.00, 1.12 |
| Partial BNT162b2 | 0.88 | 0.85, 0.91 | 0.83 | 0.81, 0.86 | 0.94 | 0.90, 0.99 |
| Full ChAdOx1 | 0.48 | 0.30, 0.78 | 0.76 | 0.70, 0.82 | 1.58 | 0.97, 2.56 |
| Full BNT162b2 | 0.32 | 0.21, 0.48 | 0.50 | 0.39, 0.65 | 1.59 | 1.07, 2.35 |
| ***Contact vaccination status*** |  |  |  |  |  |  |
| Unvaccinated | — | — | — | — | — | — |
| Partial ChAdOx1 | 0.94 | 0.91, 0.98 | 0.69 | 0.66, 0.72 | 0.73 | 0.69, 0.77 |
| Partial BNT162b2 | 0.85 | 0.82, 0.88 | 0.67 | 0.65, 0.69 | 0.79 | 0.76, 0.83 |
| Full ChAdOx1 | 0.40 | 0.27, 0.59 | 0.42 | 0.38, 0.45 | 1.04 | 0.70, 1.53 |
| Full BNT162b2 | 0.15 | 0.11, 0.21 | 0.19 | 0.16, 0.23 | 1.28 | 0.92, 1.78 |

The Delta variant was associated with increased onward transmission vs. Alpha for symptomatic index cases (aRR=1.24[1.12-1.38] when contact age=18y) and to a greater extent for asymptomatic index cases (aRR=1.40[1.22-1.59]) independent of case and contact vaccination status, but associations attenuated as contact age increased (Figure S2).

Vaccine-associated reductions in transmission post-second dose in index cases were reduced with Delta for BNT162b2 by 1.6-fold (aRR=1.59[1.07-32.35]) vs. Alpha, and likely also for ChAdOx1 (aRR=1.58[0.97-2.56]). Two BNT162b2 doses reduced transmission of Delta by more than ChAdOx1 (aRR=0.50[0.39-0.65] vs. aRR=0.76[0.70-0.82, respectively, hRR=1.51[1.15-1.97]).

### Vaccination in contacts

The estimated effect of contact vaccination status does not necessarily reflect overall vaccine effectiveness, as study inclusion was conditional on being a tested contact. However, PCR-positivity was highest in unvaccinated contacts (34,041/65,117[52%]), followed by those partially vaccinated with ChAdOx1, (3,987/12,307[32%]) and BNT162b2 (6,756/20,999[32%]) and lowest in after full ChAdOx1 (7,241/32,363[22%]) and BNT162b2 (2,642/15,457[17%]) vaccination. With Alpha, independently of effects in cases, BNT162b2-fully vaccinated contacts had lower rates of PCR-positive tests than contacts receiving ChAdOx1 (aRR 14-days post second vaccine vs. unvaccinated=0.15[0.11-0.21] vs. aRR=0.40[0.27-0.59] respectively, hRR=2.68[1.61-4.47]). With Delta, more vaccinated contacts tested PCR-positive than with Alpha, due to increases independent of vaccination status, with no strong evidence of a difference vs. Alpha in vaccine effectiveness compared to unvaccinated contacts for BNT162b2 (hRR vs. Alpha=1.26[0.91-1.75]) or ChAdOx1 (hRR=0.99[0.67-1.45]). Two doses of BNT162b2 remained more effective against Delta (aRR vs. unvaccinated=0.19[0.16-0.23]) than ChAdOx1 (aRR=0.42[0.38-0.45], hRR=2.17[1.78-2.65]).

### Duration of protection and transmission reductions

Vaccine-associated reductions in onward transmission declined over time since second vaccination in index cases (Figure 1A). Independently of contact vaccination status, for each doubling of weeks since 14 days after second vaccination in index cases, the rate of contacts testing PCR-positive increased 1.08-fold (95%CI 1.05-1.11) for ChAdOx1 and 1.13-fold (1.05-1.21) for BNT162b2 with no evidence of a difference between vaccines (hRR=0.96[0.87-1.03]). For Alpha, at two weeks post second dose of BNT162b, transmission was reduced by 68%(95%CI 52-79%) falling to 52%(29-67%) by 12 weeks, with reductions of 52%(22-70%) and 38%(−1-62%) at 2 and 12 weeks post-second ChAdOx1 dose. For Delta and BNT162b, reductions at 2 and 12 weeks were 50%(35-61%) and 24%(20-28%), respectively, and 24%(18-30%) and 2%(−2-6%) for ChAdOx1 (see Figure S5 for probabilities by case and contact vaccine status). Findings were similar restricting to contacts tested 2-7 days after the index case (Table S5, Figures S6-S7).

**Figure 1.**
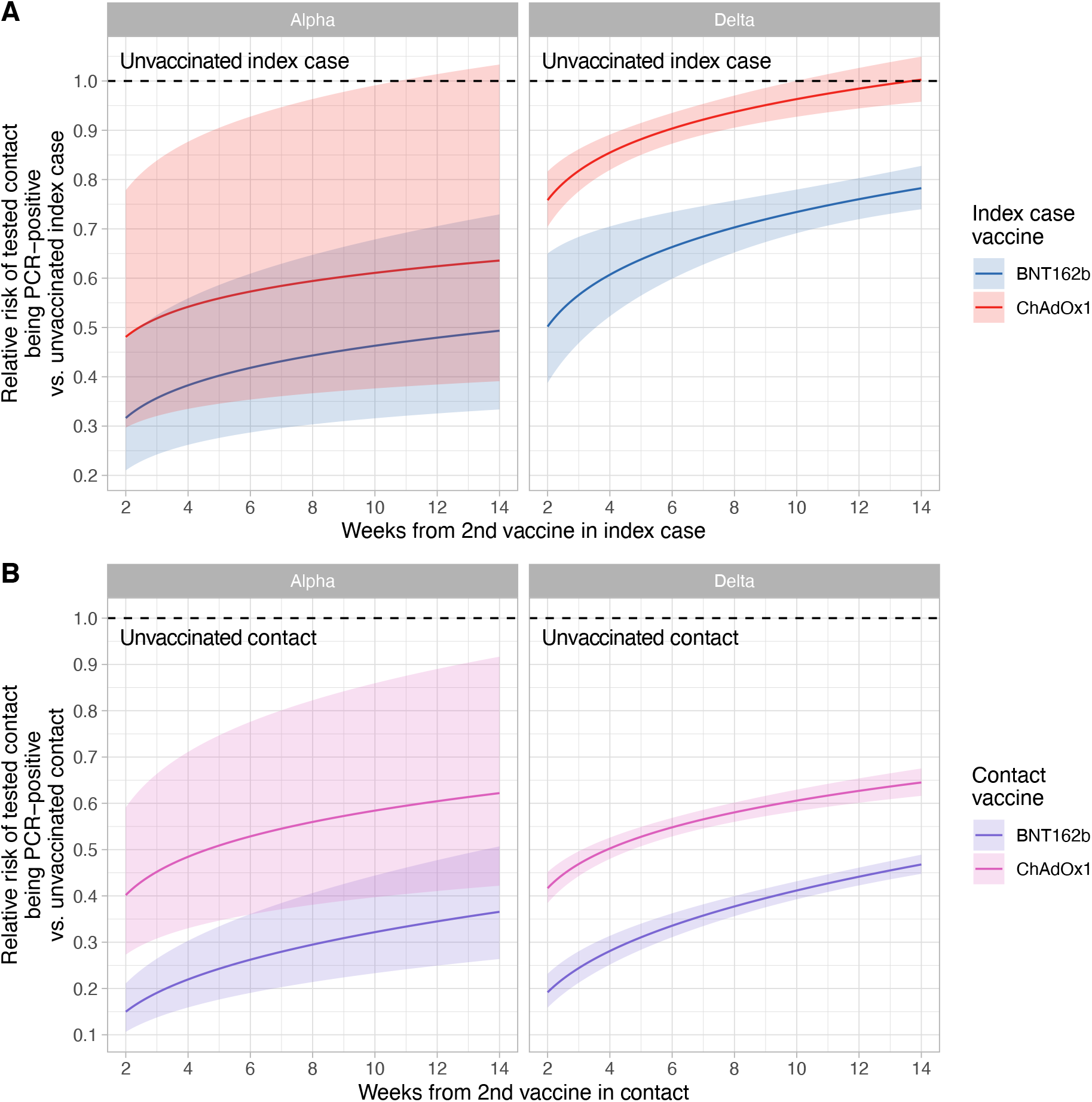
Rate ratios for positive PCR tests in contacts by time since second vaccination in index cases (panel A) and in contacts (panel B), variant, and vaccine type. Panel A compares the rate of positive PCR results in tested contacts, comparing the impact of index case vaccination status to an unvaccinated index case. Panel B compares the rate of positive PCR results in tested contacts, comparing contact vaccination status to an unvaccinated contact. See Figure S5 for probabilities of a positive test by variant and case and contact vaccination status. The shaded area indicates the 95% confidence interval. There was no evidence that fitting different rates by variant for the change in protection over weeks since second vaccine improved model fit.

Contacts receiving BNT162b2 vs. ChAdOx1 remained at lower risk of testing positive throughout 14 weeks post-second dose, despite the protective effect of BNT162b2 waning faster than ChAdOx1 (Figure 1B, aRR per doubling of weeks since 14 days after second vaccination=1.27[95%CI 1.21-1.34] vs. 1.13[1.10-1.16]; hRR=1.13[1.07-1.20]).

### Other transmission risk factors

Multiple other factors were associated with contacts testing positive (Figures 2, S2-S4, Table S4), including contact event type and index case age, with the highest rates of PCR-positivity after household contact with index cases aged ≥40 years and lower rates following contact at work/education or events/activities (Figure 2A). Contacts in their 30s and 70s had the highest rates of positive tests after household contact, while contacts in their 20s had the highest rates after contact events outside their own home (Figure 2B). Contacts of index cases of the opposite sex were more likely to test positive (Figure 2C) and male contacts were more likely than female contacts to be infected outside the home (Figure 2D).

**Figure 2.**
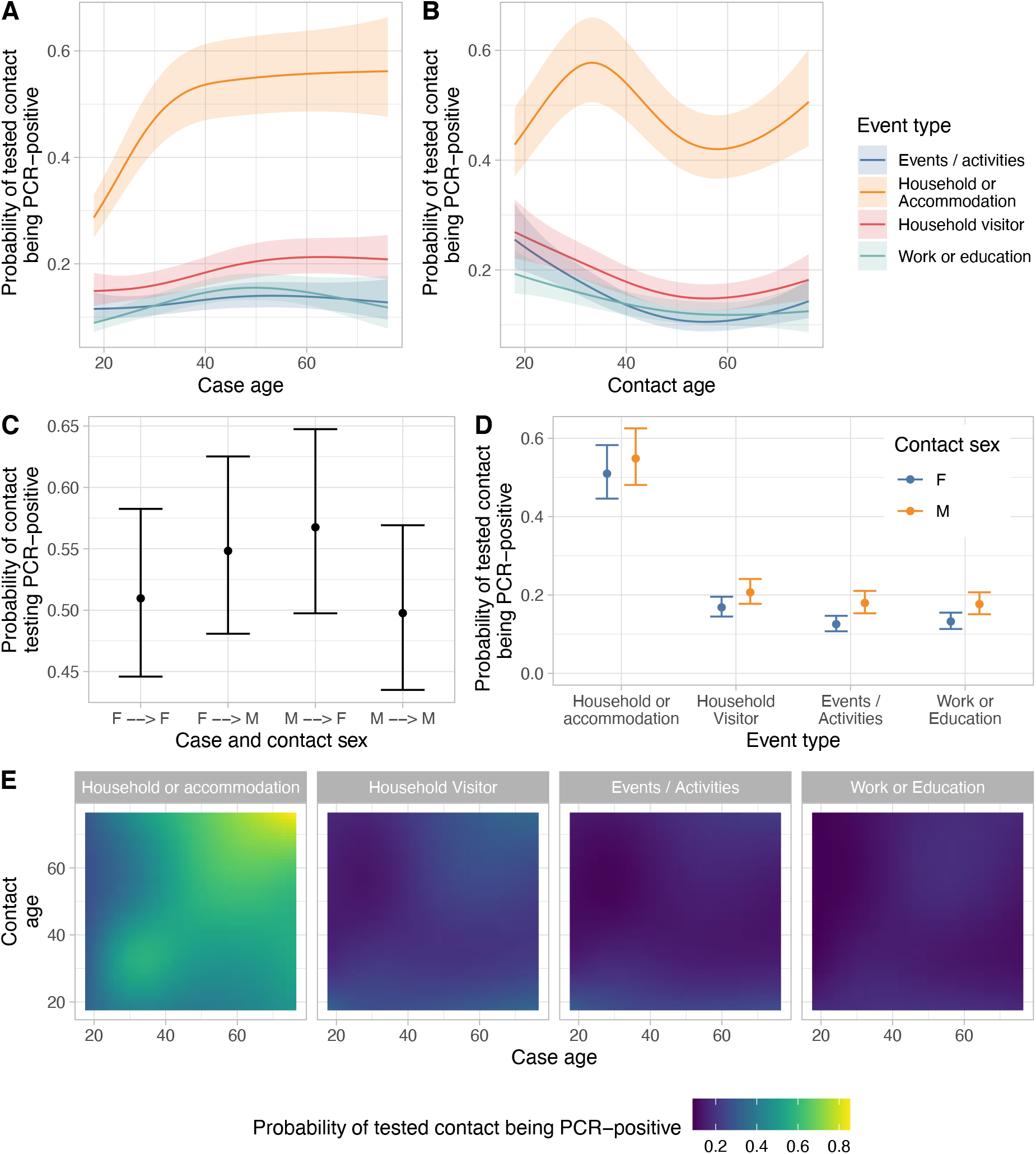
Estimated probability of a positive PCR test in contacts by contact event type and index case age (panel A) or contact age (panel B), contact sex and event type (panels C and D) and case and contact age (panel E). For each panel all other covariates are set to reference values for categorical values and median values for continuous variables, i.e. contact event type set to Household or accommodation; index case factors – age (median), sex (female), vaccination status (unvaccinated) and symptom status (symptomatic); contact factors – age (median), sex (female), vaccination status (unvaccinated); local deprivation (median), local SARS-CoV-2 incidence (median) and calendar time (median). Shaded ribbons and error bars indicate 95% confidence intervals.

Contacts of asymptomatic index cases were less likely to test positive (aRR at contact age=18y vs. symptomatic for Alpha=0.53[95%CI 0.50-0.55], Delta=0.73[0.65-0.83]) likely related to both lower viral loads (Figure 3) and lack of symptoms. Contacts living in more deprived areas and areas with higher SARS-CoV-2 incidence (Figure S3) were more likely to test positive. Positivity varied by calendar time (Figure S4).

**Figure 3.**
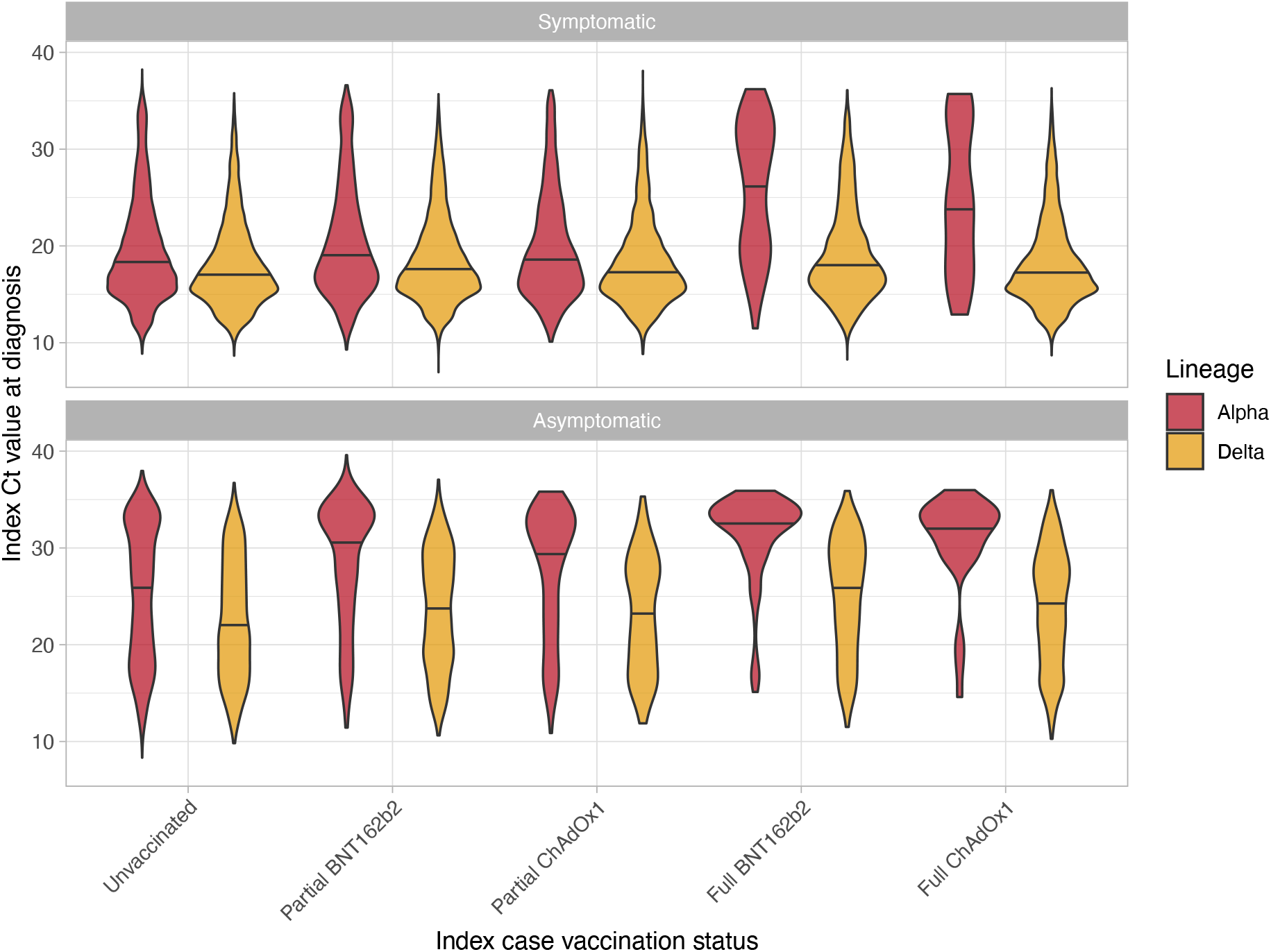
Distribution of Ct values (indicative of viral load) by index case vaccination status, variant, and symptoms. The solid line in each violin plot indicates the median. See Lee *et al* for details of equivalent viral loads in copies per ml (log_10_(VL) = 12.0 -0.328*Ct).^15^

### Extent of vaccine impact on transmission explained by Ct value

Vaccination with BNT162b2 or ChAdOx1 was associated with higher PCR Ct values (lower viral loads) in Alpha index cases at the time of their positive test, e.g. after two vaccinations with symptoms, median Ct values (IQR) were 27.4(19.7-32.1) and 23.9(18.1-32.5) vs. 18.4(15.7-22.5) if unvaccinated. However, Delta variant infections had similar Ct values whether cases were vaccinated or not (Figure 3), and lower Ct values than Alpha in both symptomatic and asymptomatic infections.

Refitting our model to include Ct values (Figure 4A), lower Ct values (higher viral loads) were independently associated with increased transmission for both Alpha and Delta, but with a greater reduction in transmission as viral load decreased (Ct increased) for Alpha vs. Delta (Figure 4B). Only a minority of the effect of full BNT162b and ChAdOx1 vaccination on transmission was mediated via variation in Ct values at the time of index case diagnosis (Figure 4C, Table S6): 18%(95%CI 9-64%) and 16%(1-80%), respectively, for Alpha and 23%(17-33%) and 7%(5-10%) for Delta.

**Figure 4.**
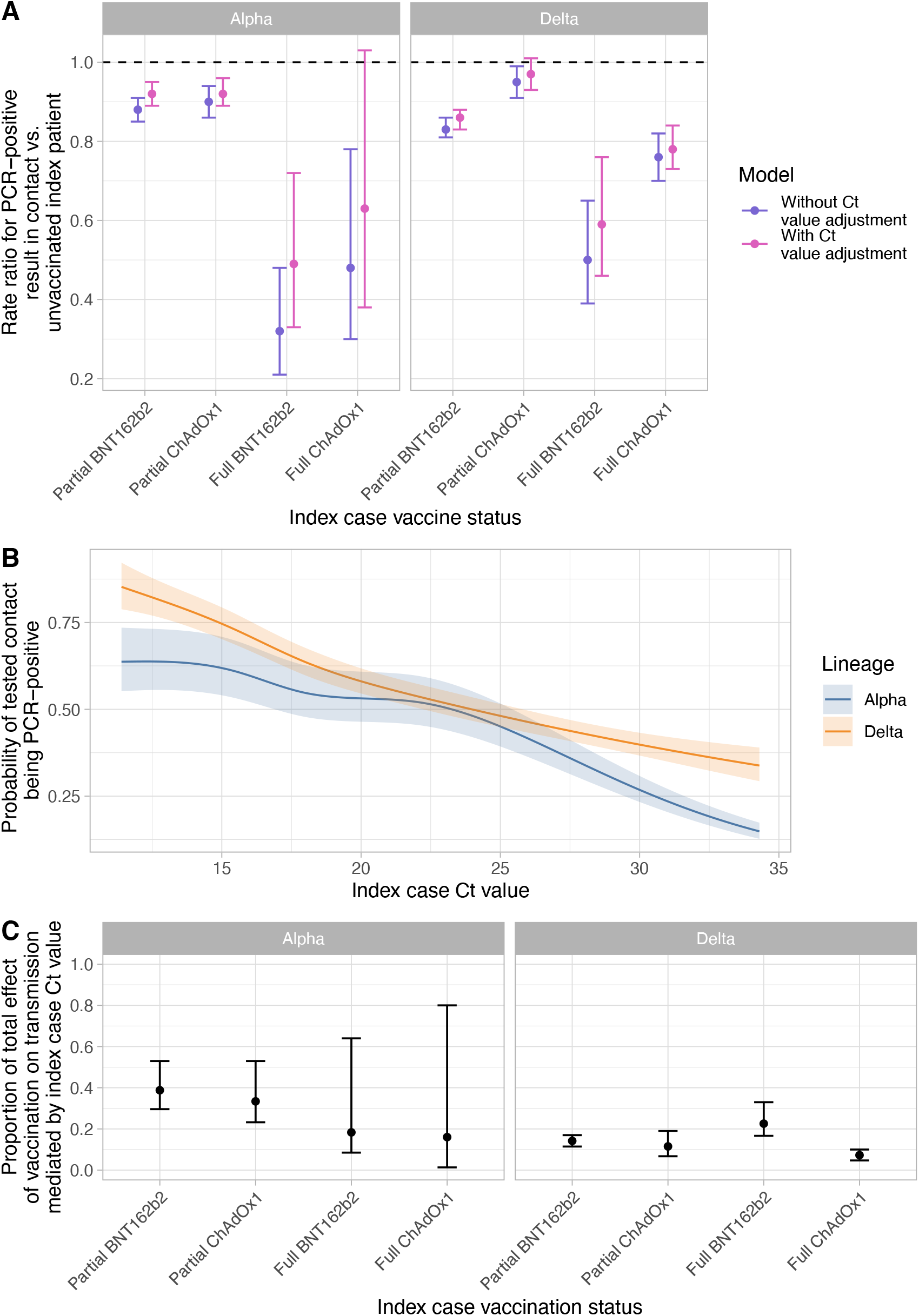
Extent of vaccine-associated transmission reductions explained by change in Ct values at index case diagnosis. Panel A shows the impact of index case vaccination on onward transmission in models with and without adjustment for index case Ct value. Panel B shows the relationship between index case Ct value and onward transmission from the model adjusting for index case Ct at the time of diagnosis. Panel C shows the proportion of the total effect of index case vaccination mediated by changes in Ct value (further details in Table S6). Adjusted rate ratios are shown in panel A and error bars (panel A and C) or the shaded ribbon (panel B) indicate the 95% confidence interval.

## Discussion

Using large-scale contact tracing data, we show that BNT162b2 and ChAdOx1 vaccination both reduce onward transmission of SARS-CoV-2 from individuals infected despite vaccination. However, reductions in transmission are lower for the Delta variant compared to Alpha for BNT162b2 and likely lower for ChAdOx1 too. Vaccines continue to provide protection against infection with Delta, but to a lesser degree than with Alpha in large population-based studies, particularly for infections with symptoms or moderate/high viral loads.^8^ Therefore, Delta erodes vaccine-associated protection against transmission by both making infection more common and increasing the likelihood of transmission from vaccinated individuals who become infected.

Vaccines have been hypothesised to reduce onward transmission from infected vaccinated individuals by reducing viral loads, as higher viral loads are associated with transmission.^14,15^However, we found that most of the effect of vaccines persisted after adjusting for Ct values at index case diagnosis; in a mediation analysis, index case Ct values only accounted for 7-23% of the impact of vaccination. This highlights that Ct values from diagnostic testing are not necessarily a surrogate of the impact of vaccination on transmission. It potentially explains why we found vaccination reduces onward Delta transmission despite Ct values being similar regardless of vaccination status. The single measured Ct value only approximates viral load at the time of transmission, as viral loads are dynamic over time.^22^ Hence, observed Ct values are likely imperfectly representative of viral loads at transmission, despite the relationship observed between Ct values at diagnosis and risk of prior onward transmission (Figure 4B). Vaccination may act by facilitating faster clearance of viable infectious virions,^17,18^ but leaving damaged ineffective virions behind that still contain PCR-detectable RNA. However this, and the performance of antigen assays post vaccination, needs further study.

We found that contacts of Delta-infected index cases vaccinated with BNT162b2 were less likely to test PCR-positive than those of index cases receiving ChAdOx1, with potentially insufficient power to resolve differences for Alpha. Contacts vaccinated twice with BNT162b2 also had lower rates of Alpha and Delta infections than those vaccinated with ChAdOx1.

Protection against onward transmission waned within 3 months post-second vaccination. For Alpha some protection against transmission remained, but for Delta this eroded much of the protection against onward transmission, particularly for ChAdOx1. “Waning” of protective behaviour may also explain some of the differences seen, with vaccination facilitating reduced social distancing and mask wearing. However, reductions in antibody levels^23^and vaccine effectiveness^8^ over time suggest biological explanations for increasing transmission are likely important. Additionally, some of the observed decline may be attributable to those clinically vulnerable with weaker immune systems being vaccinated longer ago. We also found that the probability of a contact testing positive increased with time since their second vaccination. Although BNT162b2 provided higher levels of protection for contacts throughout the 3 months post-second vaccine, protection against infection in contacts waned faster for BNT162b2 than ChAdOx1, as also seen for new infections in a representative UK survey.^8^

This study has several limitations. We considered only contacts who underwent PCR testing, to minimise bias introduced by differences in testing behaviour that may occur for multiple reasons including contacts’ vaccination status. Therefore, we cannot estimate secondary attack rates by case and contact vaccination status, and absolute protective effects of vaccination on transmission may be under-estimated as vaccine-protected uninfected contacts may not have sought testing. Our approach is also unlikely to eliminate bias, particularly if test-seeking behaviour is related to perceived vaccine efficacy, given non-specificity of many symptoms.^24^ Some contacts will have been infected by a source other than the identified ‘index case’. We restricted to contacts tested 1-10 days after an index case to minimise this, with very similar findings restricting to 2-7 days; better data on symptom onset and timing of contact events could improve estimates. We did not have sufficient data to account for previous infection status, which is also imperfectly ascertained in national testing programs. Declines over time in the adjusted probability of contacts testing positive (Figure S4) may be partly explained by increasing prevalence of prior infection in unvaccinated individuals, along with changes in test-seeking behaviour and the incidence of other infections causing similar symptoms.^25^ We used SGTF and time as a proxy for Alpha vs. Delta infection rather than sequencing, meaning some low viral load Delta infections with SGTF may have been misclassified as Alpha; however we restricted the time period of our dataset to minimise this. As we considered all PCR results in contacts, not just those tested with assays including an S-gene target, we could not assess SGTF concordance as supporting evidence for transmission between case-contact pairs. Finally, we did not have data to adjust for comorbidities; with clinically vulnerable individuals and healthcare workers vaccinated earlier and more likely to receive shorter dosing intervals. This may have impacted some findings, particularly on waning over time and differences by vaccine type; it also precluded analysis of the impact of dosing interval.^8^

The Delta variant has spread globally and caused resurgences of infection even in the setting of high vaccination coverage. Increased onward transmission from individuals who become infected despite vaccination is an important reason for its spread. Booster vaccination campaigns being considered and implemented^26^ are likely to help control transmission as well as preventing infections.

## Data Availability

Applications to use the data in this study can be made to NHS Digital's Data Access Request Service, please see https://digital.nhs.uk/services/data-access-request-service-dars for more details.

## Declarations

DWE declares lecture fees from Gilead outside the submitted work. No other author has a conflict of interest to declare.

