## Supplementary material for "The impact of SARS-CoV-2 vaccination on Alpha & Delta variant transmission"

#### Supplementary methods

##### Contact definition

Contacts were defined as follows in line with national guidelines:[1] a person who has been close to someone who has tested PCR-positive for COVID-19 anytime from 2 days before the person was symptomatic up to 10 days from onset of symptoms. The nature of the contact could include:

- Living in the same household OR
- Face to face contact (within 1 metre for any length of time) or skin to skin contact or someone the case coughed on OR
- Within 1 metre for 1 minute of longer OR
- Within 1-2 metres for more than 15 minutes OR
- Sexual contacts OR
- Travel in the same vehicle or a plane

Contacts named by more than one index patient in a 10-day period were excluded given ambiguity about the infection source if the contact tested positive.

The contact tracing data provided contained a single contact event type (household/accommodation, household visitor, events/activities, or work/education) per case-contact pair, which was determined by the original contact tracer in more specific categories which were then aggregated.

##### Vaccination status

Index cases and contacts were included where their vaccination status, obtained from National Immunisation Management Service, was known. Matching was performed using NHS numbers, which are unique to each person in the UK. As all vaccination records included an NHS number, absence of a matched record was classified as unvaccinated, accepting a small amount of misclassification from individuals vaccinated in other countries or in trials. Only individuals vaccinated with Pfizer-BioNTech BNT162b2 and AstraZeneca ChAdOx1 were included, as numbers receiving other vaccines were too few to analyse.

##### Classification of variants

Contacts of index cases tested between 01-January-2021 and 31-July-2021 were included as follows. Index cases with S-gene target failure (SGTF), used as a proxy for the Alpha (B.1.1.7) variant, were included up to 06-June-2021. During this period 5-95% of sequenced infections were due to Alpha.[2] However following 06-June-2021, the Alpha variant accounted for ≤2% of cases/week[2] (Figure S1), such that there was ambiguity about whether S gene target failure (SGTF) was due to Alpha or stochastic failure to amplify the S gene target at low viral loads with other lineages (predominantly Delta), which occurred in 1% of samples when Delta dominated at the end of July-2021. From the week beginning 10-May-2021 national spread of Delta meant that >98% of sequenced cases were either due to the Alpha or Delta variants,[2] such that we used detection of S gene on or after 10-May-2021 as a proxy for the Delta variant. Index cases without SGTF prior to 10-May-2021 were excluded, as it was not possible to distinguish which cases had the Delta variant.

#### Statistical methods details

We used multivariable Poisson regression to investigate how onward transmission, i.e., SARS-CoV-2 PCR-positive tests in contacts, varied with index case vaccination status and other variables. Poisson regression with robust standard errors[3,4] was used in place of logistic regression to improve interpretability of model outputs given a common binary outcome.

We used natural cubic splines and log transformation to account for non-linear effects of continuous variables following truncation at the 1st and 99th centiles, for splines allowing up to 5 default-placed knots (9 for calendar time to allow greater flexibility) and choosing the best fitting models using the Bayesian information criterion (BIC). Time since second vaccination for cases and contacts was $\log_2$ transformed (on the basis of model fit), truncating time separately for each vaccine. Interactions between all model main effects were included with this improved model fit based on the BIC.

We used robust standard errors with clustering based on index case identifier to account for some index cases being included in the analysis more than once if the index case had multiple contacts not named by any other source.

All analyses were performed in R, version 4.1. The sandwich library's (version 3.0) vcovCL function was used to generate variance-covariance matrices accounting for repeated measurements. Heterogeneity rate ratios and 95% confidence intervals were calculated using interaction terms and contrasts between levels of categorical variables (determined using the glht function from the multicomp library [version 1.4]).

#### Extent of vaccine-associated reductions in transmission mediated via index case Ct values at diagnosis

We refitted models including index case Ct values to investigate the relationship between Ct values (indicative of viral load[5]) and transmission. We fitted the same final model as obtained for the main analysis, and added a non-linear term spline-based term for Ct value (with up to five default placed knots as above). We tested for interactions between Ct value and all model main effects, retaining a single interaction with variant (Alpha vs. Delta) based on model fit using BIC as above.

As we find that vaccination status can impact index case Ct values, at least for Alpha variant infections, and that Ct values at index case diagnosis are associated with onward transmission, we performed a mediation analysis to assess the proportion of the total effect of index case vaccination on onward transmission that is mediated via changes in index case Ct values. To facilitate a mediation analysis using readily available methods, we fit separate models for the Alpha and Delta variants, and approximate the relationship between Ct value and onward transmission as linear (Figure 4B). Otherwise, the main outcome model remains the same. For the model representing the value of the mediator, Ct value, we use linear regression and the following covariates: index case vaccination status, time since 2$^{nd}$ index case vaccination, index case age (allowing for non-linearity), index case sex, index case symptoms and calendar time (also as a non-linear term). Analyses were performed using the mediate (version 4.5) package in R. Confidence intervals were generated using 1000 simulations and robust standard errors.

##### Sensitivity analyses – time window between case and contact PCR tests

We undertook two analyses to justify the choice of requiring contacts to be tested within 1-10 days of the index case. In the first we exploit that the likelihood of onward transmission and a PCR-positive result in a contact is approximately linearly related to the PCR Ct value recorded for the index case at diagnosis.[6] It therefore follows that where the index case is the true source, we expect this relationship to be strongest, averaging over all possible intervals from the contact event to testing of the index case. Conversely, if we fail to identify the true index case then this relationship will attenuate towards no effect. We can therefore use this property to estimate what range of time intervals between index case test and contact test that are consistent with transmission, specifically in the direction from the index case to the contact. The initial plausible range of times is determined using prior data on the serial interval between cases in a transmission chain.[7] We fitted univariable logistic regression models of the association between PCR-positive results in contacts and the Ct value in the putative index case. We fitted separate models for case-contact pairs where the contact's PCR test occurred -3, -2, -1, 0, 1, 2, 3, 4, 5, 6, 7, 8, 9, 10, 11, 12 or 13 days after the index case's PCR test. We fitted separate models for each day for Alpha and Delta variant index case infections. We then report how the odds ratio for a PCR-positive result in the contact per unit change in index case Ct value varied by time interval between case and contact tests.

In a separate sensitivity analysis we repeated the main analysis restricting to index cases and contacts where the contact was tested between 2 and 7 days following the index case.

### Supplementary results

#### Excluded index cases and contacts

A total of 27,217 contacts were excluded with incomplete data (15.7%): 16,999 contacts had no recorded vaccine status, 11 missing index case Ct values, 9,849 missing a contact event type, 182 index case sex, 5 contact sex, and 171 contact's local area. The remaining 146,243 case-contact pairs had complete data for all study covariates.

#### Relationship between index case Ct value and onward transmission, by time between index case and contact PCR tests

The most common time interval between index case and contact PCR tests was 0 days, with most other contacts tested in the 7 days following the index case (Figure S5A).

The odds ratio for a PCR-positive test in the contact per unit change in index Ct value varied by the time interval between case and contact PCR tests (Figure S5B). For Alpha variant infections, the odds ratio was greatest for contact PCR tests done 1 to 10 days after the index case. For Delta, the odds ratio was greatest for tests 1 to 6 days after the index case. This suggests that these time intervals are most enriched for index cases that are the true source for the infection in their contact. Whereas, for 'contacts' tested before index cases the odds ratio attenuated to near 1, i.e. no effect, suggestive that the contacts acquired their infection from another source (some may even have been the source of the 'index case's' infection). As relatively few contacts were tested >7 days after the index case, the upper limit set on the time interval between tests is less critical.

#### Sensitivity analysis restricting to contacts tested 2-7 days after an index case

Findings were similar to the main analysis. A table analogous to Table 1 for the sensitivity analysis is shown as Table S5 and a figure analogous to Figure 1 as Figure S6.

#### Supplementary Tables

| Model factors | Description | | Notes |
|---|---|---|---|
| Main exposures | Index patient vaccination status | | Pre-specified interaction with index patient SARS-CoV-2 variant included |
| | Index patient SARS-CoV-2 variant | | Alpha; Delta |
| Covariates | Contact event type | | |
| | Index patient factors | Age | |
| | | Sex | |
| | | Symptom status | Symptoms were determined at the point of contact tracing, which followed a positive PCR test in the index case. Therefore, individuals who were symptomatic or pre-symptomatic at the contact event are classed as symptomatic. Those who had no symptoms up to the point of contact tracing are recorded as asymptomatic. |
| | Contact factors | Age | |
| | | Sex | |
| | | Vaccination status | Pre-specified interaction with index patient SARS-CoV-2 variant included |
| | Local area deprivation | | Index of multiple deprivation (IMD) score for the local area (lower tier local authority, LTLA) containing the contact's home address. These data were obtained from publicly available national statistics: https://www.gov.uk/government/statistics/english-indices-of-deprivation-2019. |
| | Local area SARS-CoV-2 Incidence | | Rolling 7-day average SARS-CoV-2 incidence in the contact's LTLA on the day on the contact's PCR test |
| | Local area second vaccine uptake | | The percentage of those eligible having received two vaccines in the contact's LTLA on the day of the contact's PCR test |
| | Calendar time | | |

**Table S1. Model covariates**.

| Index case vaccination status | n | Age, median | Age, IQR | Female, n | Female, % | Symptomatic, n | Symptomatic, % | Alpha variant, n | Alpha variant, % |
|---|---|---|---|---|---|---|---|---|---|
| Unvaccinated | 59,956 | 35 | 25 - 50 | 30,691 | 51.2 | 56,184 | 93.7 | 43167 | 59,956 |
| Partial ChAdOx1 | 8,294 | 45 | 40 - 52 | 4,345 | 52.4 | 7,670 | 92.5 | 3024 | 8,294 |
| Partial BNT162b2 | 20,927 | 28 | 22 - 35.5 | 10,107 | 48.3 | 19,634 | 93.8 | 3256 | 20,927 |
| Fully ChAdOx1 | 15,086 | 49 | 36 - 57 | 7,590 | 50.3 | 14,182 | 94 | 67 | 15,086 |
| Fully BNT162b2 | 4,235 | 48 | 32 - 60 | 2,621 | 61.9 | 3,754 | 88.6 | 127 | 4,235 |

**Table S2. Index case demographics, symptom status, and infecting variant by index case vaccine status**.

| Contact vaccination status | n | Age, median | Age, IQR | Female, n | Female, % | Alpha variant, n | Alpha variant, % |
|---|---|---|---|---|---|---|---|
| Unvaccinated | 65,117 | 37 | 26 - 51 | 34,654 | 53.2 | 52,321 | 80.3 |
| Partial ChAdOx1 | 12,307 | 47 | 41 - 54 | 7,026 | 57.1 | 3,739 | 30.4 |
| Partial BNT162b2 | 20,999 | 30 | 24 - 37 | 12,010 | 57.2 | 3,829 | 18.2 |
| Fully ChAdOx1 | 32,363 | 53 | 45 - 58 | 18,888 | 58.4 | 151 | 0.5 |
| Fully BNT162b2 | 15,457 | 51 | 38 - 60 | 10,628 | 68.8 | 337 | 2.2 |

**Table S3. Contact demographics, and associated index case variant by contact vaccine status.**

| **Characteristic** | **PCR tested contacts** | **PCR-positive contacts** | **% infected** | **aRR** | **95% CI** | **p-value** |
|---|---|---|---|---|---|---|
| Variant | | | | | | |
| Alpha | 60,377 | 31,326 | 52 | — | — | |
| Delta | 85,866 | 23,341 | 27 | 1.24 | 1.12, 1.38 | <0.001 |
| Index case vaccination status | | | | | | |
| Unvaccinated | 76,401 | 35,459 | 46 | — | — | |
| Partial ChAdOx1 | 11,236 | 3,878 | 35 | 0.90 | 0.86, 0.94 | <0.001 |
| Partial BNT162b2 | 31,039 | 7,947 | 26 | 0.88 | 0.85, 0.91 | <0.001 |
| Full ChAdOx1 | 21,421 | 6,067 | 28 | 0.48 | 0.30, 0.78 | 0.003 |
| Full BNT162b2 | 6,146 | 1,316 | 21 | 0.32 | 0.21, 0.48 | <0.001 |
| Index case vaccination status * Variant | | | | | | |
| Partial ChAdOx1 * Delta | 7,617 | 2,236 | 29 | 1.06 | 1.00, 1.12 | 0.046 |
| Partial BNT162b2 * Delta | 27,122 | 6,162 | 23 | 0.94 | 0.90, 0.99 | 0.017 |
| Full ChAdOx1 * Delta | 21,322 | 6,053 | 28 | 1.58 | 0.97, 2.56 | 0.065 |
| Full BNT162b2 * Delta | 5,970 | 1,294 | 22 | 1.59 | 1.07, 2.35 | 0.021 |
| Index case, per doubling of weeks since two weeks after second ChAdOx1 dose | | | | 1.08 | 1.05, 1.11 | <0.001 |
| Index case, per doubling of weeks since two weeks after second BNT162b2 dose | | | | 1.13 | 1.05, 1.21 | 0.001 |
| Index case age | See Figure 2A | | | | | |
| Index case sex | | | | | | |
| F | 75,678 | 27,182 | 36 | See Figure 2C | | |
| M | 70,565 | 27,485 | 39 | | | |
| Index case symptoms | | | | | | |
| Symptomatic | 136,534 | 52,583 | 39 | — | — | |
| Asymptomatic | 9,709 | 2,084 | 21 | 0.53 | 0.50, 0.55 | <0.001 |
| Variant * Index case symptoms | | | | | | |
| Delta * Asymptomatic | 4,834 | 779 | 16 | 1.12 | 1.04, 1.22 | 0.005 |
| Event type | | | | | | |
| Household or accommodation | 97,204 | 46,437 | 48 | See Figures 2A, 2B | | |
| Household Visitor | 16,505 | 3,284 | 20 | | | |
| Events / Activities | 16,114 | 2,560 | 16 | | | |
| Work or Education | 16,420 | 2,386 | 15 | | | |
| Contact vaccination status | | | | | | |
| Unvaccinated | 65,117 | 34,041 | 52 | — | — | |
| Partial ChAdOx1 | 12,307 | 3,987 | 32 | 0.94 | 0.91, 0.98 | 0.004 |
| Partial BNT162b2 | 20,999 | 6,756 | 32 | 0.85 | 0.82, 0.88 | <0.001 |
| Full ChAdOx1 | 32,363 | 7,241 | 22 | 0.40 | 0.27, 0.59 | <0.001 |
| Full BNT162b2 | 15,457 | 2,642 | 17 | 0.15 | 0.11, 0.21 | <0.001 |

| Characteristic | PCR tested contacts | PCR-positive contacts | % infected | aRR | 95% CI | p-value |
|---|---|---|---|---|---|---|
| Contact vaccination status * Variant | | | | | | |
| Partial ChAdOx1 * Delta | 8,568 | 2,299 | 27 | 0.73 | 0.69, 0.77 | <0.001 |
| Partial BNT162b2 * Delta | 17,170 | 5,040 | 29 | 0.79 | 0.76, 0.83 | <0.001 |
| Full ChAdOx1 * Delta | 32,212 | 7,221 | 22 | 1.04 | 0.70, 1.53 | 0.85 |
| Full BNT162b2 * Delta | 15,120 | 2,611 | 17 | 1.28 | 0.92, 1.78 | 0.14 |
| Contact, per doubling of weeks since two weeks after second ChAdOx1 dose | | | | 1.13 | 1.10, 1.16 | <0.001 |
| Contact, per doubling of weeks since two weeks after second BNT162b2 dose | | | | 1.27 | 1.21, 1.34 | <0.001 |
| Contact age | See Figure 2B | | | | | |
| Contact sex | | | | | | |
| F | 83,206 | 30,047 | 36 | See Figures 2C, 2D and S2 | | |
| M | 63,037 | 24,620 | 39 | | | |
| Index of multiple deprivation, per 1000 change, higher indicates more deprived | | | | 1.01 | 1.01, 1.01 | <0.001 |
| Local SARS-CoV-2 incidence | See Figure S3 | | | | | |
| Study day | See Figure S4 | | | | | |
| Event type * Contact sex | See Figure S2 | | | | | |
| Index case sex * Contact age | See Figure 2D | | | | | |
| Index case sex * Contact sex | See Figure 2C | | | | | |
| Event type * Index case age | See Figure 2A | | | | | |
| Event type * Contact age | See Figure 2B | | | | | |
| Index case age * Contact age | See Figure 2E | | | | | |

**Table S4. Multivariable logistic regression analysis of associations of PCR-positive results in contacts**. aRR, adjusted rate ratio; CI, confidence interval; * indicates an interaction term. Non-linear relationships and associated interactions are shown in Figures 2 and S2-S4 as indicated. There is an interaction between variant and contact age, such that all rate ratios for variant are shown at a contact age of 18 years.

| | Alpha | | Delta | | Delta vs. Alpha | |
|---|---|---|---|---|---|---|
| **Characteristic** | **aOR** | **95% CI** | **aOR** | **95% CI** | **Interaction OR** | **95% CI** |
| ***Impact on onward transmission: Case vaccination status*** | | | | | | |
| Unvaccinated | — | — | — | — | — | — |
| Partial ChAdOx1 | 0.89 | 0.85, 0.94 | 0.95 | 0.91, 1.00 | 1.07 | 1.00, 1.14 |
| Partial BNT162b2 | 0.88 | 0.84, 0.92 | 0.84 | 0.81, 0.87 | 0.96 | 0.91, 1.01 |
| Full ChAdOx1 | 0.48 | 0.25, 0.93 | 0.78 | 0.71, 0.85 | 1.60 | 0.83, 3.07 |
| Full BNT162b2 | 0.30 | 0.18, 0.49 | 0.48 | 0.35, 0.66 | 1.62 | 1.03, 2.58 |
| ***Contact vaccination status*** | | | | | | |
| Unvaccinated | — | — | — | — | — | — |
| Partial ChAdOx1 | 0.98 | 0.94, 1.03 | 0.73 | 0.70, 0.77 | 0.75 | 0.70, 0.80 |
| Partial BNT162b2 | 0.89 | 0.86, 0.93 | 0.70 | 0.68, 0.73 | 0.79 | 0.75, 0.83 |
| Full ChAdOx1 | 0.45 | 0.29, 0.69 | 0.46 | 0.42, 0.50 | 1.02 | 0.66, 1.56 |
| Full BNT162b2 | 0.15 | 0.09, 0.22 | 0.21 | 0.17, 0.27 | 1.51 | 0.98, 2.34 |

**Table S5. Sensitivity analysis: Relationship between PCR-positive results in contacts, and index case and contact vaccination status according to Alpha/Delta variant in the index case (restricting to contacts tested 2 to 7 days inclusive after an index case)**. Results for those with two vaccine doses are estimated at day 14 post second vaccine, see Figure S7 for trends with time post-second vaccine. aRR, adjusted rate ratio, CI confidence interval. Adjustment made for contact event type; index case factors - age, sex, and symptom status; contact factors - age, sex; local deprivation, local SARS-CoV-2 incidence and calendar time.

| Index case vaccine status | Variant | Total effect | Average causal mediation effect, via Ct value | Average direct effect, not via Ct value | Proportion mediated |
|---|---|---|---|---|---|
| Full BNT162b2 | Alpha | -0.37 (-0.48, -0.15) | -0.07 (-0.12, -0.04) | -0.30 (-0.43, -0.04) | 0.18 (0.09, 0.64) |
| Full BNT162b2 | Delta | -0.17 (-0.21, -0.13) | -0.04 (-0.05, -0.03) | -0.13 (-0.18, -0.08) | 0.23 (0.17, 0.33) |
| Full ChAdOx1 | Alpha | -0.37 (-0.50, -0.03) | -0.07 (-0.12, -0.02) | -0.30 (-0.45, 0.07) | 0.16 (0.01, 0.80) |
| Full ChAdOx1 | Delta | -0.10 (-0.12, -0.08) | -0.01 (-0.01, 0.00) | -0.09 (-0.11, -0.07) | 0.07 (0.05, 0.10) |
| Partial BNT162b2 | Alpha | -0.06 (-0.08, -0.04) | -0.02 (-0.03, -0.02) | -0.04 (-0.05, -0.02) | 0.39 (0.30, 0.53) |
| Partial BNT162b2 | Delta | -0.06 (-0.06, -0.05) | -0.01 (-0.01, -0.01) | -0.05 (-0.06, -0.04) | 0.14 (0.11, 0.17) |
| Partial ChAdOx1 | Alpha | -0.05 (-0.07, -0.03) | -0.02 (-0.02, -0.01) | -0.03 (-0.05, -0.02) | 0.33 (0.23, 0.53) |
| Partial ChAdOx1 | Delta | -0.04 (-0.05, -0.03) | 0.00 (-0.01, 0.00) | -0.03 (-0.05, -0.02) | 0.12 (0.07, 0.19) |

**Table S6. Extent of vaccine-associated reductions in transmission mediated via index case Ct values at diagnosis**. The reference group for each comparison is unvaccinated index cases. Effects reported are averaged over the dataset, i.e. both vaccinated and unvaccinated contacts and all times since second vaccination in index cases. Because times since second vaccination to infection were typically longer for BNT162b2 than ChAdOx1, the average total effect for BNT162b is more similar to ChAdOx1 than is seen in Figure 1 where a representative range of times since second vaccination are shown.

#### Supplementary Figures

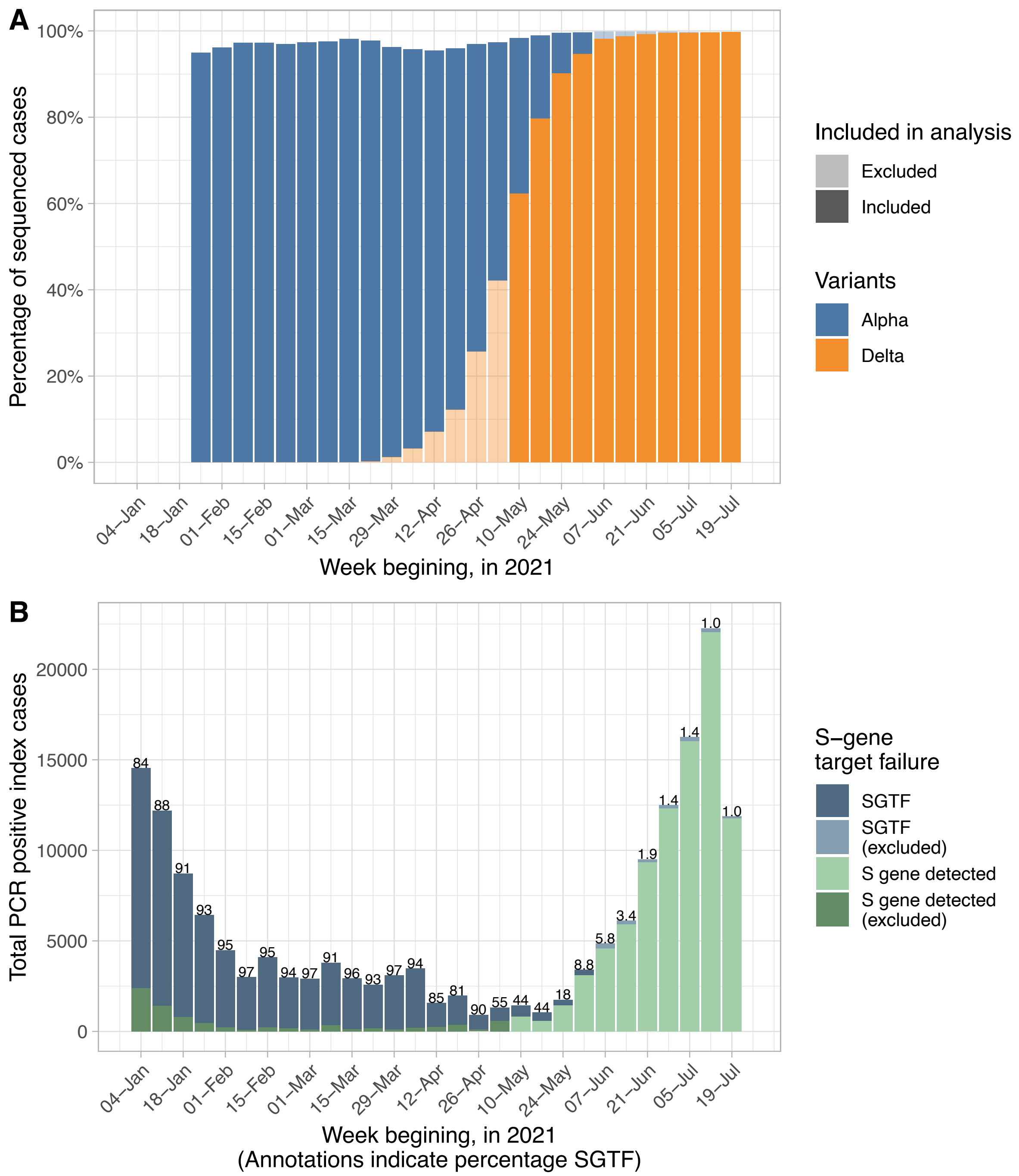


**Figure S1. National incidence of Alpha and Delta variants as determined by whole genome sequencing (panel A) and index case incidence according to S-gene target failure (SGTF, panel B)**. In panel A, weeks for which index cases with presumed Alpha and Delta infections (see Methods) were included in the analysis are shown in the denser shading (sequencing not used to determine variant as not available for all index cases). Based on results of national sequencing data available at https://www.gov.uk/government/publications/investigation-of-novel-sars-cov-2-variant-variant-of-concern-20201201. SGTF was seen in 1% of infections in the final two weeks of the study when Delta accounted for nearly all infections. Hence by only classifying index cases Alpha variant on the basis of SGTF in weeks where Alpha prevalence by sequencing exceeded ≥5% we ensured misclassification was minimised. In panel B the percentage of index cases with SGTF is provided above each bar.

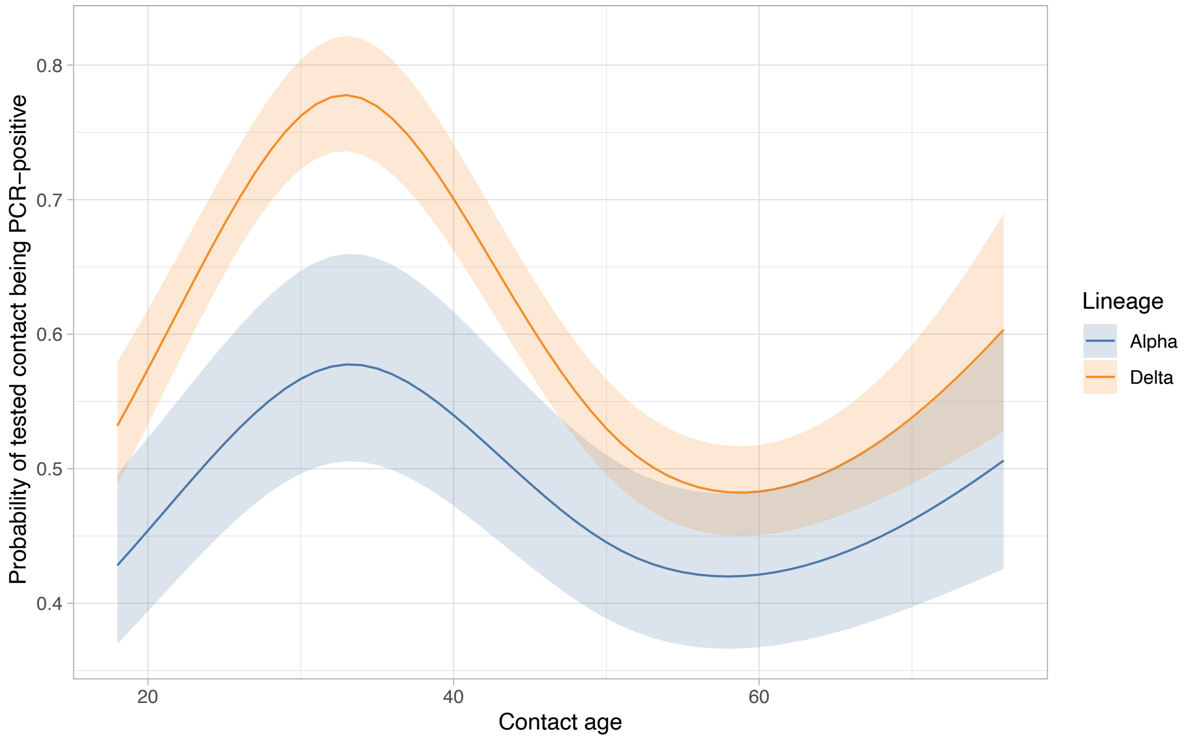


**Figure S2. Relationship between contact age and variant and probability of a positive PCR test in a contact**. All other continuous covariates are set to median values and categorical covariates to reference categories. The error bars indicate 95% confidence intervals.

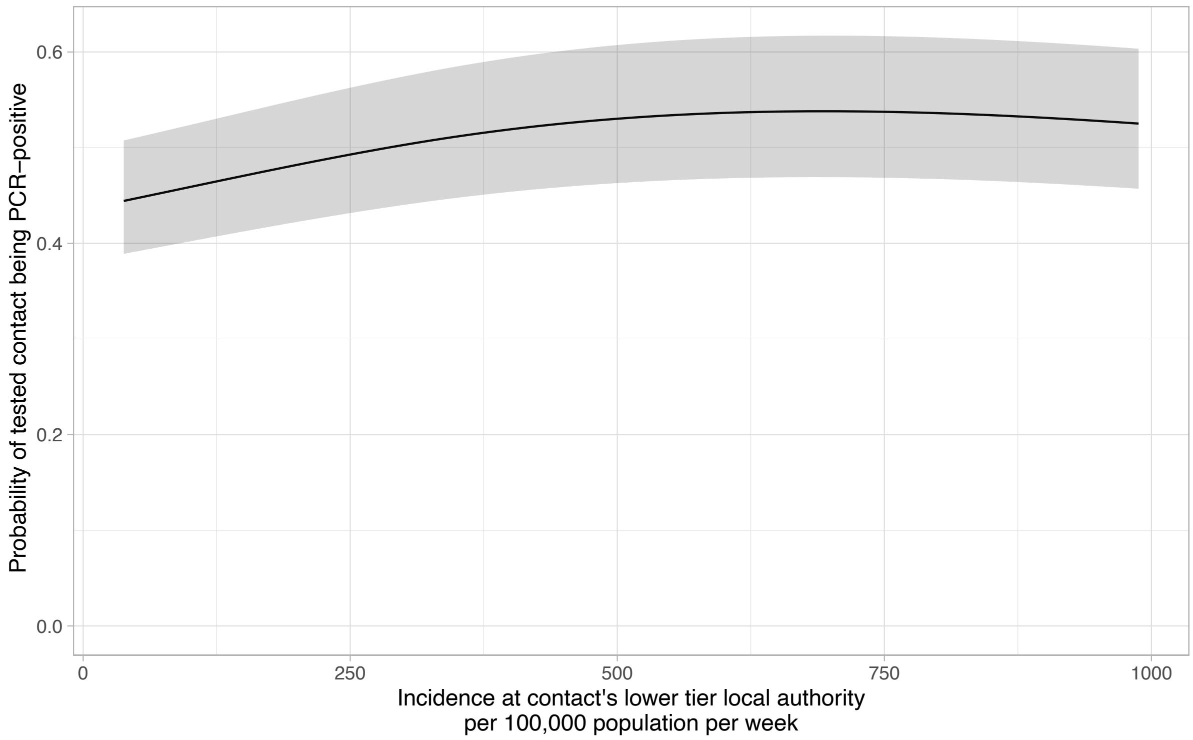


**Figure S3. Relationship between local SARS-CoV-2 incidence and probability of a positive PCR test in a contact**. All other continuous covariates are set to median values and categorical covariates to reference categories. The shaded area indicates the 95% confidence interval.

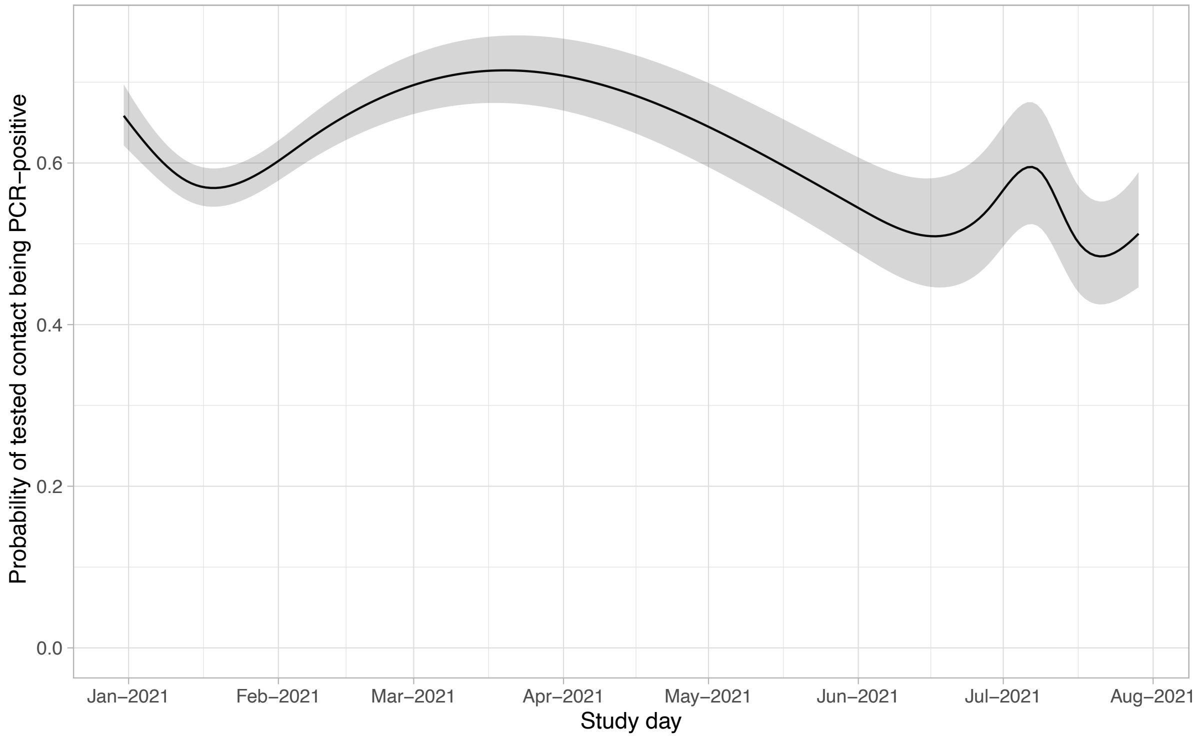


**Figure S4. Relationship between study day and probability of a positive PCR test in a contact**. All other continuous covariates are set to median values and categorical covariates to reference categories. The shaded area indicates the 95% confidence interval. Note “UEFA Euro 2020” European Football Championship held between 11 June and 11 July 2021.

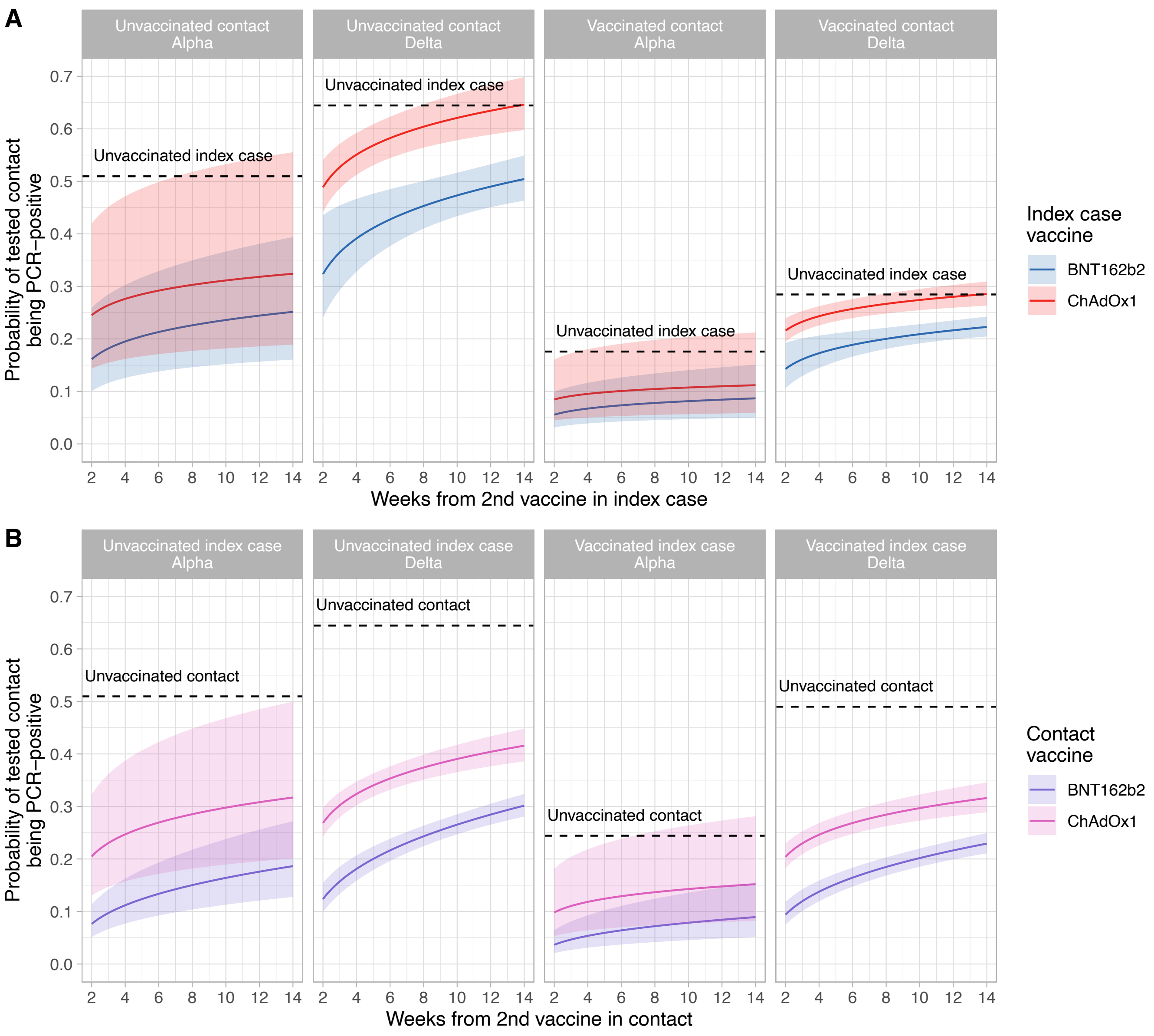


**Figure S5. Estimated probability of a positive PCR test in contacts by time since second vaccination in index cases (panel A) and in contacts (panel B), variant, and vaccine type**. For panel A estimates are displayed for an unvaccinated contact and a fully BNT162b vaccinated contact at 12 weeks post second dose. For panel B estimates are shown for an unvaccinated index case a fully BNT162b vaccinated index case at 12 weeks post second dose. The dashed horizontal lines indicate the probability of a positive-PCR result in an unvaccinated contact of an unvaccinated index case. The shaded area indicates the 95% confidence interval. Adjustment made for covariates, set to reference values: contact event type (set to Household or accommodation); index case factors – age (median), sex (female), and symptom status (symptomatic); contact factors – age (median), sex (female); local deprivation (median), local SARS-CoV-2 incidence (median) and calendar time (median). Note only 26.2% of contacts had a PCR test, as testing was predominantly only performed if symptoms developed. Therefore, overall secondary attack rates are lower than shown here restricting only to contacts undergoing testing.

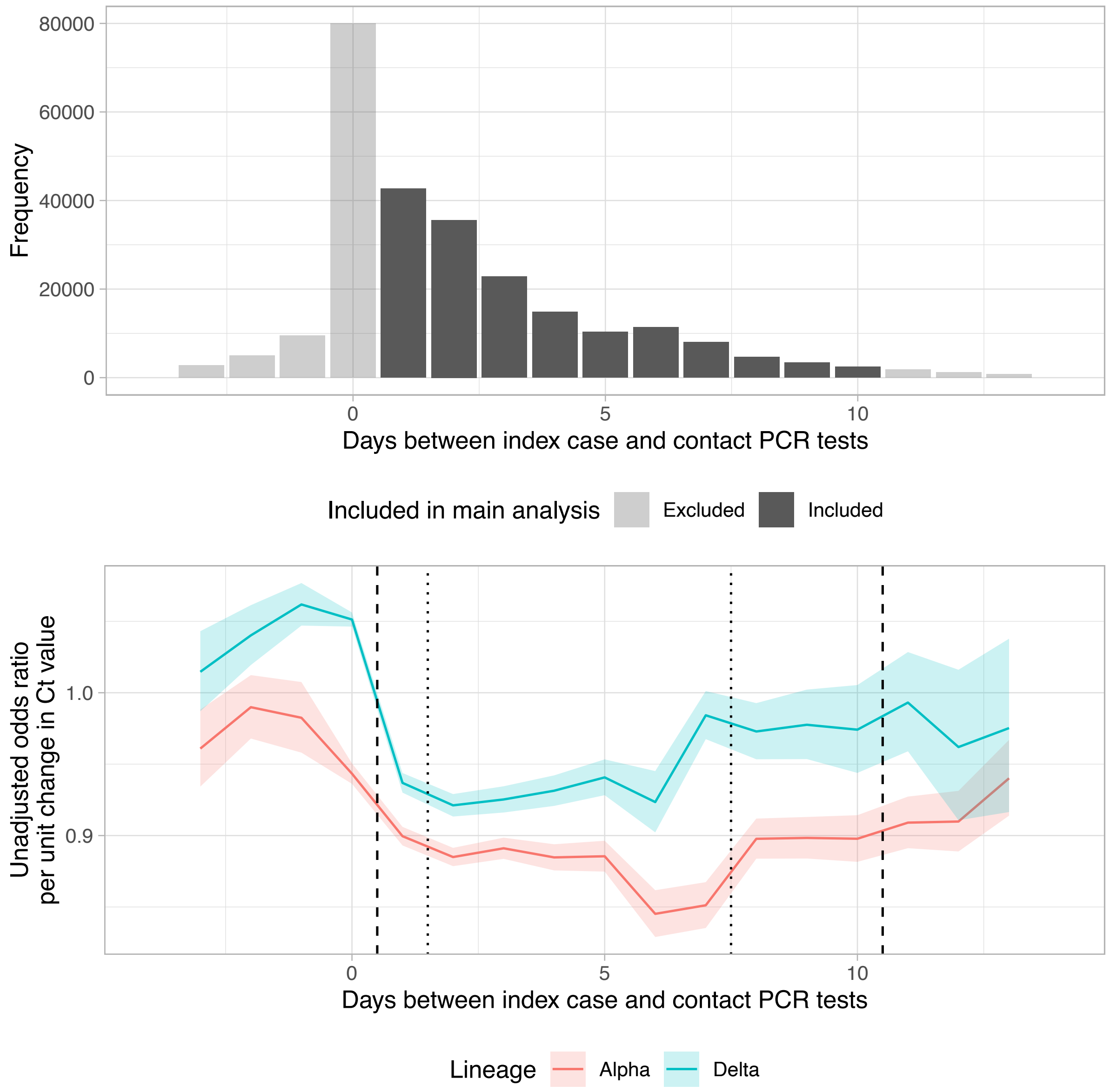


**Figure S6. Relationship between index case Ct value and onward transmission, by time between index case and contact PCR tests**. Panel A shows the distribution of days between index case and contact PCR tests in case-contact pairs, shaded according to which pairs were included in the main analysis. Panel B shows the univariable odds ratio for a positive-PCR result in a contact for each unit change in index case Ct value, according to days between index case test and contact test and SARS-CoV-2 variant. The dashed vertical lines indicate the cut-offs for inclusion in the main analysis (days 1-10 inclusive) and the dotted vertical lines the cut-offs for inclusion in a separate sensitivity analysis (days 2-7 inclusive).

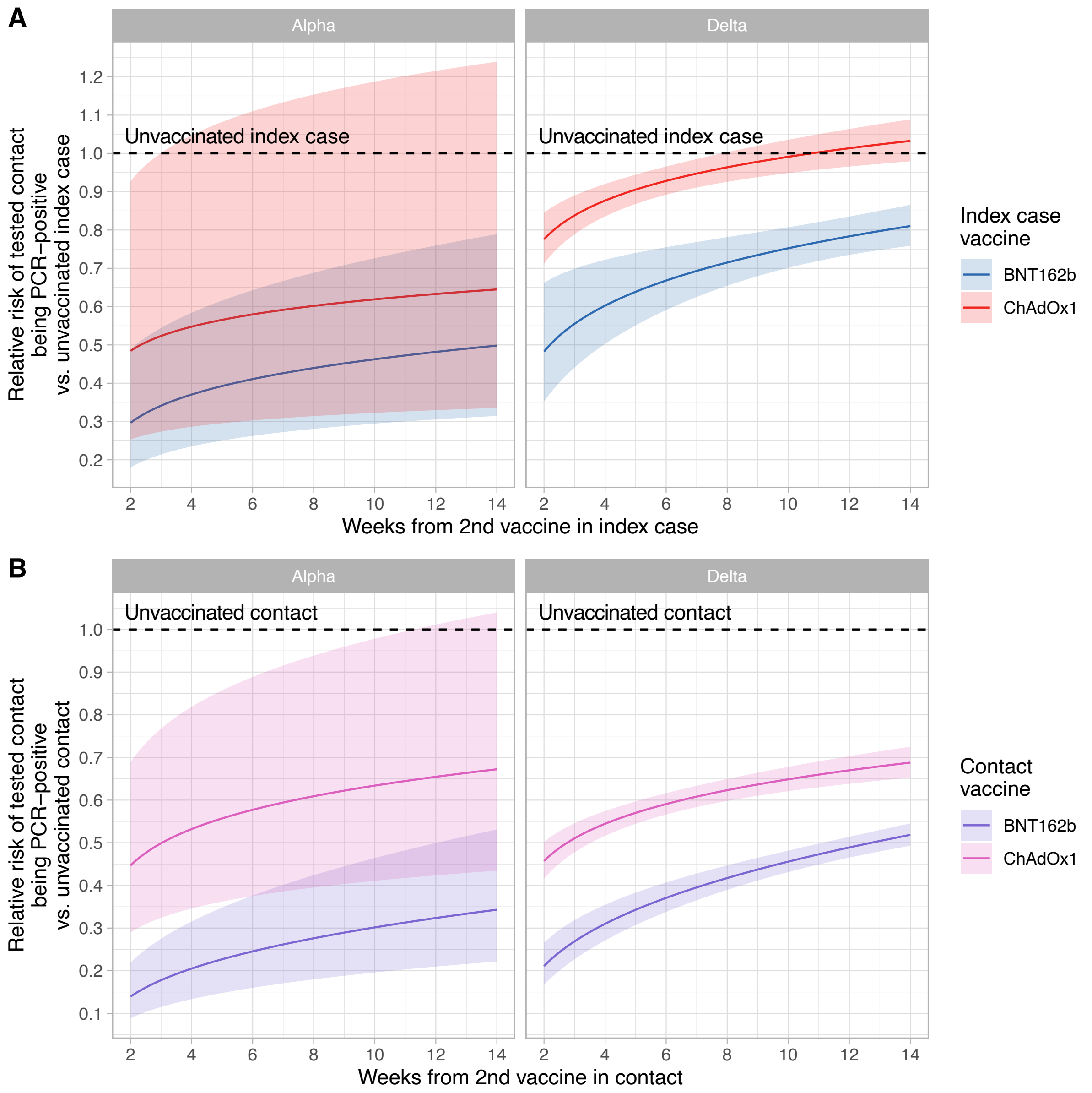


**Figure S7. Sensitivity analysis: Rate ratios for positive PCR tests in contacts by time since second vaccination in index cases (panel A) and in contacts (panel B), variant, and vaccine type (restricting to contacts tested 2 to 7 days inclusive after an index case)**. Panel A compares the rate of positive PCR results in test contacts, comparing index case vaccination status to an unvaccinated index case. Panel B compares the rate of positive PCR results in test contacts, comparing contact vaccination status to an unvaccinated contact. The shaded area indicates the 95% confidence interval.
